# Disruptions in Care: Consequences of the COVID-19 Pandemic in a Children’s Hospital

**DOI:** 10.1101/2021.12.02.21266778

**Authors:** Catherine Diskin, Julia Orkin, Blossom Dharmaraj, Tanvi Agarwal, Arpita Parmar, Kelly Mc Naughton, Eyal Cohen, Alia Sunderji, David Faraoni, Annie Fecteau, Jason Fischer, Jason Maynes, Sanjay Mahant, Jeremy N Friedman

## Abstract

**Background:** Public health restrictions are an essential strategy to prevent the spread of COVID-19; however, unintended consequences of these interventions may have led to significant delays, deferrals and disruptions in medical care. This study explores clinical cases where the care of children was perceived to have been negatively impacted as a result of public health measures and changes in healthcare delivery and access due to the COVID-19 pandemic.

**Methods:** This study used a qualitative multiple case study design with descriptive thematic analysis of clinician-reported consequences of the COVID-19 pandemic on care provided at a children’s hospital. A quantitative analysis of overall hospital activity data during the study period was performed.

**Results:** The COVID-19 pandemic has resulted in significant change to hospital activity at our tertiary care hospital, including an initial reduction in Emergency Department attendance by 38% and an increase in ambulatory virtual care from 4% before COVID-19, to 67% in August, 2020. Two hundred and twelve clinicians reported a total of 116 unique cases. Themes including (1) timeliness of care, (2) disruption of patient-centered care, (3) new pressures in the provision of safe and efficient care and (4) inequity in the experience of the COVID-19 pandemic emerged, each impacting patients, their families and healthcare providers.

**Conclusion:** Being aware of the breadth of the impact of the COVID-19 pandemic across all of the identified themes is important to enable the delivery of timely, safe, high-quality, family-centred pediatric care moving forward.

**What’s new:** COVID-19 disrupted typical paediatric care delivery.

This study demonstrates the breadth of its’ impact on the delivery of timely, safe, equitable and patient and family centered care, highlighting considerations for paediatric providers as we move forward.

## Introduction

Since the onset of the COVID-19 pandemic, much of society, including healthcare delivery, has changed.^1^ In March 2020, healthcare services underwent a major reorganization,^2^ with non-essential activity including in-person ambulatory activity and elective surgery curtailed, guided by a provincial directive.^3^ Clinicians raised concerns regarding the potential impact on morbidity and mortality of patients experiencing illness during the pandemic. Delayed presentation to care, deferral of care, impact on cancer treatment and disruption of clinical pathways to accommodate COVID-19 have been described.^4,5^ Children have been affected by multiple consequences that have spanned their physical, social developmental and emotional wellbeing.^6,7^ Early data from Italy highlighted a sharp reduction in children presenting to acute care^6^ and significant morbidity, including presumed preventable intensive care admissions due to children presenting late in their course of illness.^6,8^ Although important for reducing viral transmission, the potential risk of school closure and societal lockdown to children has led to a call for urgent monitoring and systematically collected data.^9, 10, 11,^

We hypothesized that the COVID-19 pandemic and efforts undertaken in the hospital sector to mitigate the effects may have unintended secondary consequences. This study’s primary objective is to describe courses of care for hospitalized children that were altered by the COVID-19 pandemic from the clinician’s perspective. We planned to identify thematic similarities to inform clinical practice and explore the associated negative effects of health care and hospital policy changes associated with the COVID-19 pandemic.

## Methods

This mixed-methods study was performed at the Hospital for Sick Children, a 350-bed tertiary-care children’s hospital in Toronto, Canada, and describes the experience between March and August 2020, which coincides with the first wave of the COVID-19 pandemic in Canada.^12^

### Study Design

This prospective study involved a qualitative multiple case study design with descriptive thematic analysis of clinician-reported consequences of the COVID-19 pandemic on care provided at a children’s hospital. Study data was collected and managed using the Research Electronic Data Capture (REDCap) platform. REDCap is a secure, web-based software platform designed to support data capture for research studies.^13,14^ A quantitative review of hospital data was also performed to understand and contextualize patterns of clinical activity changes during the study period. The study protocol was reviewed and approved by the Hospital for Sick Children’s Research Ethics Board (#1000070386). A brief interim report, completed mid-way through the study, was previously published.^15^ We disseminated early results in real-time to inform healthcare leaders and decision-makers about the breadth of the impact associated with the COVID-19 pandemic.

### Methodology

#### (1) Case series

Cases were identified in two ways in order to facilitate comprehensive capture; 1) a prospective bi-weekly email survey, and 2) monthly review of clinical cases submitted for morbidity and mortality (M&M) review hospital-wide. The review of cases submitted as part of the M&M process was intended to enhance the comprehensiveness of data.

##### a) Clinician reporting

A bi-weekly survey was sent to all physicians (including trainees), dentists and advanced practice nurses (n=1727) from May 25 to August 25, 2020. The survey included demographic information as well as case identification questions. They were asked to identify any patients they perceived to have experienced a suboptimal quality of care or health outcome related to changes that had occurred as a result of the COVID-19 pandemic, including their perception of the impact (Supplementary appendix – Table 1).

Individuals could complete the survey on more than one occasion if they experienced other cases that met the criteria.

##### b) M&M records

Two reviewers considered written reports of hospital-wide morbidity and mortality (M+M) meetings until December 31, 2020, to identify any additional cases that listed the COVID-19 pandemic as contributing to the reported morbidity. New cases that were identified within the M+M reporting structure (i.e., those not already identified by clinicians) were included in the same database as those identified by the clinician-survey and subsequent case study analysis.

### Case Study Analysis

Analysis followed a qualitative case series methodology using a narrative synthesis approach to determine similarities and associated themes.^16^ Data were extracted from the hospital record for all reported clinical cases (Supplementary appendix - Table 2) focussing on the morbidity experienced. Three independent research team members (TA, CD and JO) undertook the thematic analysis. This involved (1) data familiarization, (2) data coding, (3) consideration of themes, (4) revision of themes, and (5) analysis of individual themes.^17^ A pattern that emerged from the dataset as key to understanding the study question was identified as a theme.^17^

Several overarching themes emerged, and data were grouped into clusters to characterize and situate the data.^17^ Themes were reviewed and defined such that the analytic narrative and data extracts are weaved together and contextualized within real-life context^16^, within the existing literature.^17^ When reviewers disagreed, cases were discussed including a review of objective evidence, until a consensus was reached. Some cases reflected multiple themes.

#### (2) Hospital Utilization & Clinical Activity

Using the institutions’ decision support analytics, hospital activity data was obtained to understand changes in clinical care activity, including presentations to the Emergency Department (ED), hospital admissions, surgeries, and radiological tests. Hospital activity data was compared between March-August 2019 to March-August 2020.

## Results

### Case series

#### Survey participants

Two hundred and twelve clinicians from all hospital departments (Pediatrics, Perioperative Services, Diagnostic Imaging, Psychiatry and Laboratory Medicine) completed at least one survey during the study period (Table 1). Twenty clinical sub-specialties within the Department of Pediatrics and 10 in Perioperative Services were represented (Supplementary appendix - Table 3). One hundred and sixteen cases were reported (some respondents completed the survey and did not report a case). Four cases were previously reported, and nine cases did not have sufficient detail to guide a case review. One case reported as a delayed acute presentation was excluded, as on review, symptoms were present for less than 24 hours. A review of M&M data identified 3 cases where the pandemic was listed as a contributory factor, two of which were already reported by survey respondents.

**Table 1:**
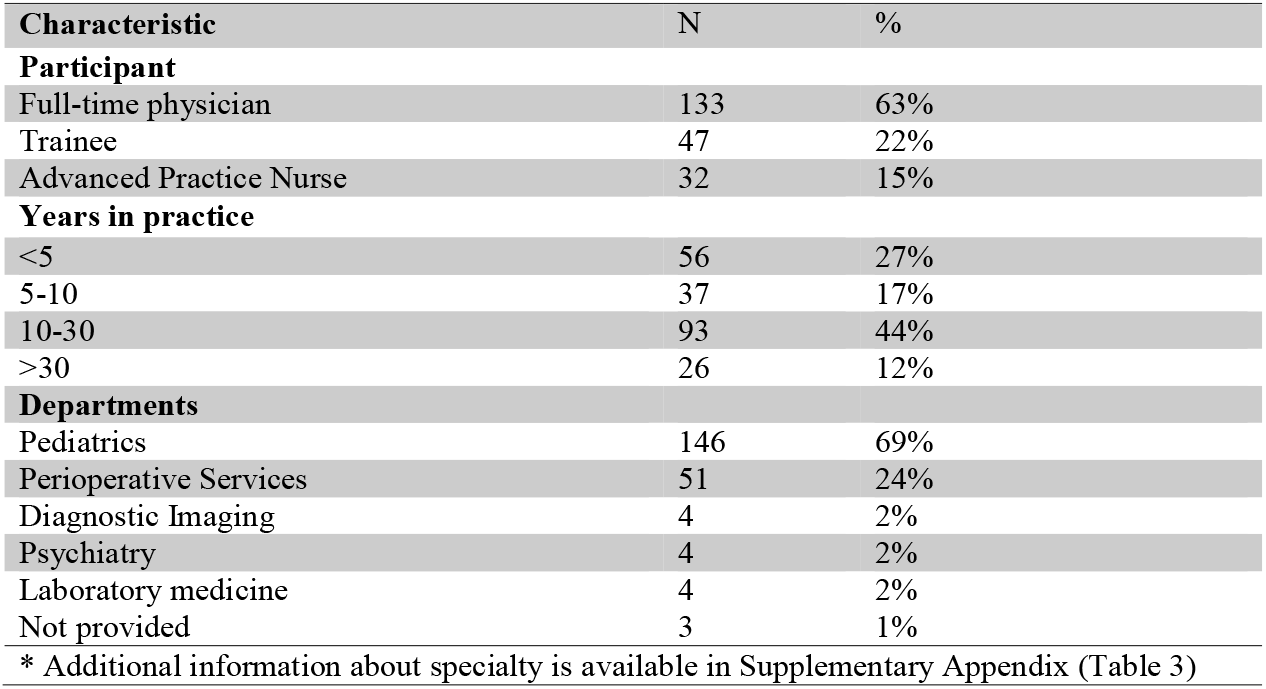
Clinician demographics*.

| **Characteristic** | **N** | **%** |
| --- | --- | --- |
| **Participant** |  |  |
| Full-time physician | 133 | 63% |
| Trainee | 47 | 22% |
| Advanced Practice Nurse | 32 | 15% |
| **Years in practice** |  |  |
| <5 | 56 | 27% |
| 5-10 | 37 | 17% |
| 10-30 | 93 | 44% |
| >30 | 26 | 12% |
| **Departments** |  |  |
| Pediatrics | 146 | 69% |
| Perioperative Services | 51 | 24% |
| Diagnostic Imaging | 4 | 2% |
| Psychiatry | 4 | 2% |
| Laboratory medicine | 4 | 2% |
| Not provided | 3 | 1% |

\* Additional information about specialty is available in Supplementary Appendix (Table 3)

#### Thematic analysis

Several broad themes emerged, including (1) timeliness of care, (2) disruption of patient-centered care, (3) inequity in the experience of the COVID-19 pandemic and (4) new pressures in the provision of safe and efficient care. Within each of these themes, subthemes emerged, highlighting the impact on (1) patients, (2) their families and (3) healthcare providers.

#### (1) Timeliness of care (68 cases)

Clinicians reported that secondary to the COVID-19 pandemic, delayed acute presentation (n=24), postponement of scheduled procedures (n=22), caregiver cancellation of ambulatory clinical appointments (n=13) and disruption of community care (n=9) impacted the care of their patients.

##### Impact on children

Clinicians reported parents describing deferral and delay in accessing acute medical care because of concerns of COVID-19 exposure. For example, a clinician reported a perceived delay in presentation due to family reluctance to attend healthcare in the care of an adolescent later diagnosed with Burkitt’s lymphoma presenting with a 5-week-history of dyspnea and dysphagia. Parents also rescheduled planned care, including surgical procedures and ambulatory care, due to fear of exposure to COVID-19. Clinicians highlighted that the deferral of scheduled clinical activity was associated with consequences. Examples ranged from the inability to remove port-a-caths quickly, resulting in a perceived increased risk of central line-associated bloodstream infections, to a child’s loss of motor skills and increased pain while awaiting orthopedic intervention for hip subluxation.

Further, cases reported challenges beyond the hospital, including access to community services. Rehabilitation services, including access to physiotherapy and occupational therapy, were reduced. Also, concerns regarding community services such as the newborn hearing screening program being suspended, leading to concerns regarding potential harm.

##### Impact on families

Clinicians described how families adapted their decision-making, e.g., when to seek medical care. Some decided to wait based on their ability to reach their primary care provider. Clinicians also described their perception of increased caregiver burden related to deferred interventions such as orthopedic surgical care whereby children/youth experienced increasing pain necessitating additional services including referrals to the chronic pain service.

##### Impact on clinicians

In addition to providing clinical care and supporting patients and families to navigate the healthcare system during a pandemic, clinicians adjusted their decision-making during a time of enforced restricted activity, e.g., reviewing and re-prioritizing previously planned interventions including diagnostic imaging investigations, interventional radiology procedures and surgeries. They reported distress prioritizing cases in ways they never had to do before. Clinicians managed the ramp-up of clinical activity, e.g., resumption of surgical activity, with many additional restrictions and polices in place such as infection control procedures and/or enhanced environmental decontamination procedures, contributing to a stressful experience.

#### (2) Disruption to the delivery of patient and family-centered care (18 cases)

Eighteen cases described the impact on the hospital or clinicians’ ability to provide patient and family-centered care; 9 related directly to the child’s experience and 9 to the family experience.

##### Impact on children

As part of routine screening for COVID-19, multiple nasopharyngeal swab tests mandated by hospital policy (e.g., screening before a procedure or on admission) was a reported cause of distress to patients. Some children described by survey respondents had four swabs performed in less than three weeks. Visitor policy restrictions were put in place at the hospital to limit presence and decrease risk of infectious spread; and clinicians reported related challenges e.g., at the end of a child’s life, when siblings and extended family were not present.

##### Impact on families

Clinicians highlighted the broader impact of limiting family presence on the entire family. The lack of two caregivers was often described as causing additional distress amongst both children and parents alike. Conversations involving the disclosure of important information, e.g., providing a new serious diagnosis, often involved one parent present and another joining remotely, causing distress amongst the family as reported by clinicians. Also, the family policy restriction limited family caregivers to only one caregiver at the bedside, which for children with complex needs was sometimes inadequate. Lastly, the usual supports, including parental overnight accommodation, were closed, and alternatives, e.g., hotels, were expensive, which clinicians reported increased stress and burden on families.

##### Impact on clinicians

Clinicians described how they were required to enforce policies to miminize infectious spread that conflicted with their ability to provide optimal patient and family-centered care. An example of this was the enforcement of the reduced family presence at the bedside policy. Clinicians described experiencing moral distress when caring for a child at end of life where family presence, including siblings, remained limited.

#### (3) Equity (17 cases)

##### Impact on children

Clinicians described the impact of school closures on children, specifically those who require specialized services within the school setting, such as those with developmental disabilities requiring specialized therapy. For example, one case reported a young person with autism presenting with increased anxiety and skill regression perceived to be associated with loss of resources due to school closure, resulting in increased medication and additional support required for the family. Another report described the challenges a family experienced supporting their child with autism participate in virtual learning, ultimately opting for home-schooling, thereby creating additional stress in the home.

Clinicians reported disruption to services designed to ensure child wellbeing, e.g., child protection services were limited in their ability to complete in-home visits for child protection concerns on account of inadequate personal protective equipment (PPE) availability. The provision of virtual care highlighted various issues relating to equity explored when discussing safe and effective care (next section).

##### Impact on families

Clinicians reported missed or cancelled appointments as parents, especially those who were essential workers unable to work from home, struggled to find adequate support due to the closure of daycare and/or school and decreased availability of family members to care for siblings because of social distancing measures. Also, the hospital closed its child-minding services, including supervised space for siblings to play while their family attends an appointment. Some families opted to defer appointments, investigations or treatment, including essential services and treatments such as chemotherapy for ongoing cancer care.

##### Impact on clinicians

Clinicians described their frustration and stress due to the limited ability to respond to the inequities they witnessed while trying to help families navigate diminished supports, e.g., access to low-cost accommodation during their child’s prolonged hospital stay remote from their home.

#### (4) Safe and effective care (21 cases)

##### Impact on children

The safe and effective care of patients was impacted by the previously described themes, including timeliness of care and equity. Additional challenges to providing effective care are illustrated by a case that reflected the difficulties in establishing a therapeutic relationship through virtual care with a young person with severe anxiety. Another case described a child who required admission to the hospital to complete imaging investigations for a headache. This would typically be completed in the Emergency Department; however, admission was required as COVID-19 test results were necessary before the provision of general anesthesia.

##### Impact on families

Clinicians reported increased challenges related to complex discharge planning. For example, discharge home from hospital for a child with medical complexity and multiple technology dependencies (ventilator-dependent, tracheostomy, enterostomy feeds) was delayed due to lack of community homecare supports. This was coupled with parental hesitancy to receive home care services related to the risk of COVID-19 exposure.

Ten cases reported were related to the provision of virtual care, with staff and families experiencing challenges relating to communication, e.g., providing laboratory requisitions was more challenging with many families not owning a printer. Staff described that physical signs were missed, e.g., pleural effusion, contributing to delay in arriving at correct diagnosis and treatment provision. In addition, examples of communication challenges related to not being face-to-face contributing to error, e.g., incorrect dosage of medication being taken, were reported.

##### Impact on clinicians

Clinicians reported difficulties in adapting to care provision in an environment of rapid change with new, frequently updated policies. The requirement for the use of PPE, including a mask and eye protection for all patient encounters was reported as a distraction. One case reported fogging of eyewear to have contributed to the incorrect reading of a medication pump and additional communication challenges were reported including the reduced opportunity to read non-verbal cues.

One case reported that the absence of team members on in-patient ward rounds, a result of efforts to reduce gatherings on the ward, impacted team performance, e.g., the pharmacist’s absence on the ward round reduced the opportunity to identify medication-related errors.

A clinician described increased diagnostic anchoring with a tendency toward COVID-19 related diagnoses. For example, a child presenting with extremity changes, swelling and redness of the right foot was misdiagnosed as having ‘COVID toes,’ and the correct diagnosis of arterial thrombus was initially missed, compounded by the virtual nature of the physical examination.

### Hospital activity (March-August 2020)

Attendance in the ED (Figure 1) decreased by 39%, from 36,940 to 22,542 visits between March and August 2020 compared with the same period in 2019. The percentage of patients seen in the ED requiring admission increased from 10.5% to 15.3% during this period.

**Figure 1:**
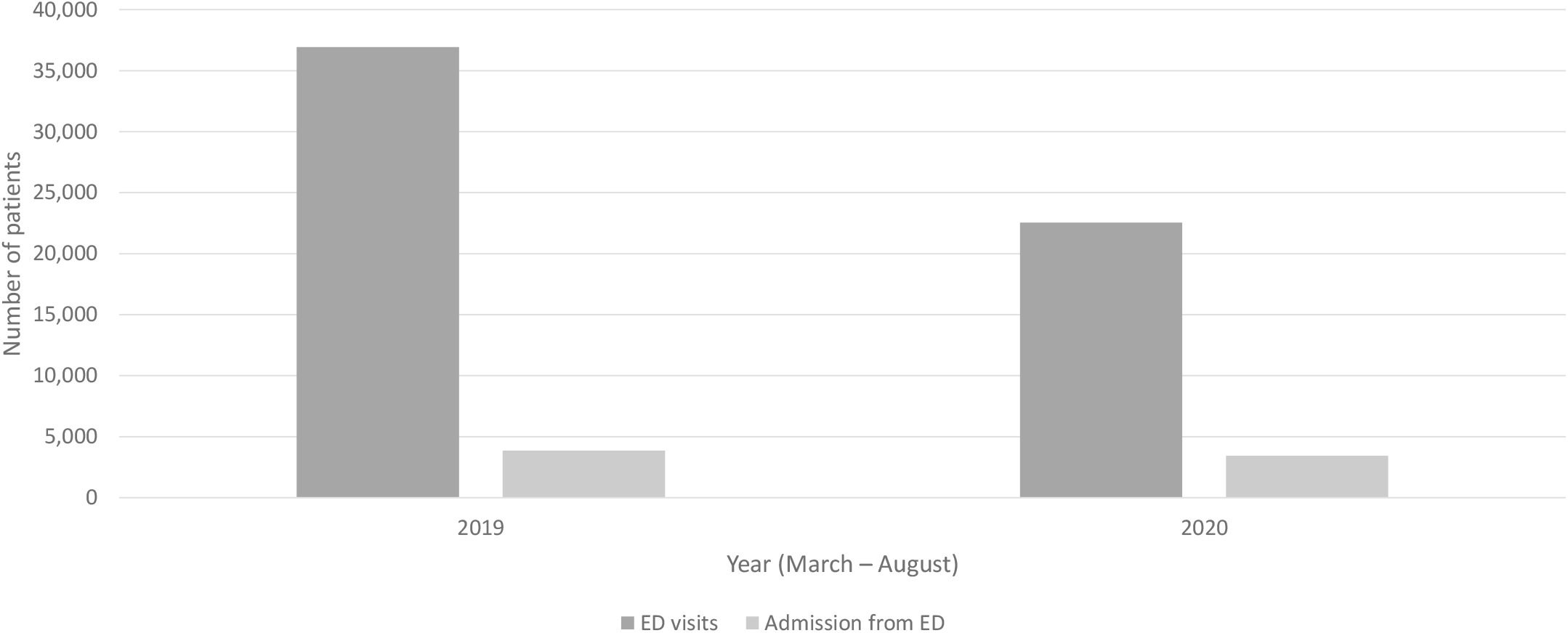
Emergency Department activity at SickKids (March to August 2019 versus March to August 2020)

From April to August 2020, there was a 17% reduction in surgeries and 33% reduction in outpatient radiological investigations compared with the same period in 2019. The number of children awaiting surgical intervention in August 2020 had increased by 31% since the start of the COVID-19 pandemic. The pandemic triggered an increase in virtual care, particularly in ambulatory care. Virtual visits increased from 4% of all ambulatory visits before COVID-19, to 67% of all ambulatory visits in August, 2020.

## Discussion

The study results explore the broad impact of the COVID-19 pandemic on pediatric hospital care from the perspective of a large group of clinicians. Delays in presentation for care during the pandemic, and the potential impact on morbidity, including hospitalization and financial cost, have been previously described. ^18,19,20^ Our hospital data confirm a drop in ED attendance as well as reductions in surgical and ambulatory activity, and a large switch to virtual care in the ambulatory setting. Our results illustrate that the impact of the COVID-19 pandemic extend beyond simply access to timely care to much broader health quality domains including patient-centeredness, equity and safety. Our findings highlight three key areas of concern specifically relating to patient and family-centered care, the expansion of virtual care and care of vulnerable populations.

Family-centered care, a standard of care in many institutions caring for children, involves taking a partnered healthcare decision-making approach.^21^ The provision of patient and family-centered care during the pandemic was challenged. The core tenant of shared-decision making was often limited due to policies in place including family presence at the bedside.^22^ Particular situations, e.g. when providing a new diagnosis require careful consideration as family presence can support parental coping and mitigate decisional conflict ^23, 24^. Hospitals need to continue to learn from their growing experience of providing healthcare during a pandemic and balance policies to align with the best care, including family-centeredness, an essential contributor to patient and family wellbeing.^25^ Innovative interventions could support healthcare providers to engage with families, e.g. using technology to support sibling involvement at the bedside.

Clinicians reported the challenges they encountered as they adapted to virtual care delivery in a rapidly changing environment, echoing previous experience that emphasized integrating virtual care with existing systems.^26^ They were often unable to provide care as they previously did, e.g., not completing a physical examination or reduced ability to read non-verbal communication cues. This highlights that the delivery of virtual care requires a particular skillset on the part of the healthcare provider including decision-making about the appropriateness of virtual care.^27^

The expansion of virtual care is associated with benefit, possibly enhancing family and patient-centered care delivery. For example, virtual ambulatory care can reduce the frequency with which families have to travel and attend hospital, reducing the need for caregivers to take time off work and associated costs. Coordinating the involvement of multiple professionals in a clinical interaction is potentially easier, with many able to join virtually. As a potentially valuable means to support patient-centered care, virtual care needs to be championed.^26^

However, as healthcare providers, we need to be aware that inequities in digital health exist alongside other factors contributing to poorer health outcomes such as poverty. Lack of access to technology or the expertise to navigate it can contribute to health inequities associated with increased age, lower level of educational attainment and lower socio-economic status.^28^ Advances in virtual care need to be accompanied by a concerted effort to prevent disparities in care for patients without access to internet or devices,^29^ including alternatives for families unable to attend virtual appointments.

Our findings support previously published commentaries and research studies, highlighting subgroups of children as particularly vulnerable, including those with medical complexity, developmental disabilities and mental health diagnoses.^30–33^ As families continue to provide care to their children with additional needs during the pandemic,^34^ we need to consider children who are particularly vulnerable to its impact, e.g., those who receive healthcare and therapy via the educational system. Clinicians must continue to advocate for paid sick leave and other policies which support and facilitate family caregiver’s interactions with healthcare.

Clinicians reported increasing moral distress and burnout throughout the COVID-19 pandemic.^35^ Cases reported highlighted the challenges clinicians face as witnesses of the inequity within society and healthcare. In addition, clinicians themselves are also likely experiencing similar issues such as reduced childcare availability, school closures, and sick loved ones. It is important to remind leaders and managers in healthcare to be mindful of the burden that healthcare professionals are currently bearing, particularly as the pandemic stretches on. Understanding the link between clinicians, family and patient experience, the role of societal and institutional policies, and actions undertaken at various levels in response to lived experience and policies is an important area of study, both to support healthcare workers and deliver family and patient-centered care.

### Limitations

This study was performed in a single tertiary-care pediatric academic centre, limiting its generalizability. We recognize that the approach taken by hospitals to the COVID-19 pandemic may vary.^36^ The case finding methodology used was not real-time and cannot provide a denominator or frequency for events. However, a thorough review of M&M records uncovered only 1 additional case, suggesting that the frequency of survey distribution and its prospective nature might mitigate this limitation. The study is clinician-centric, but involved only doctors and advanced practice nurses. A broader representation of health care providers including bedside nurses and allied health professionals would result in a richer understanding of the disruption in care related to the COVID-19 pandemic. The involvement of patients and family caregivers is required to enhance our understanding^37^, as the challenges faced by patients due to delays might be overlooked. More subtle manifestations of inequity may have been overlooked as the study did not examine the various contributing factors to the individual experience of the pandemic. Lastly, the results reflect the experience of frontline clinicians who chose to respond and are therefore subject to their bias. To truly understand the parent and child perspective, we plan to further engage with families and describe their experiences as a future step in this work.

## Conclusion

The broad consequences of health system changes as a result of the COVID-19 pandemic have impacted patients, families, healthcare providers and the healthcare system as a whole. Understanding the breadth of this impact is essential as we strive to deliver safe, high-quality, family-centered pediatric care in this new era. As the pandemic continues, we need to carefully consider how best to provide elective and ambulatory care, including surgery, in this era of infection control. Particular attention should be paid to ensuring timely access to safe care for children with special needs and families from disadvantaged settings lacking in resources, as well as the impact of the COVID-19 pandemic on the frontline clinician.

## Data Availability

All data produced in the present study are available upon reasonable request to the authors

## Contributor Statements

## Acknowledgments (if any)

The authors acknowledge our colleagues’ support at The Hospital for Sick Children, including Ms. Ethel Lagman, Kate Langrish and Karima Karmali.

## Abbreviations used

M+M: morbidity and mortality
PPE: personal protective equipment

## References

1. Miller GA, Buck CR, Kang CS, et al. COVID-19 in Seattle - Early Lessons Learned. J Am Coll Emerg Physicians Open. 53(9):1689–1699. doi:10.1002/emp2.12064

2. Willan J, King AJ, Jeffery K, Bienz N. Challenges for NHS Hospitals during Covid-19 Epidemic. BMJ. 2020;368(March):1–2. doi:10.1136/bmj.m1117

3. Ministry of Health Ontario. COVID-19 Operational Requirements: Health Sector Restart. 2020;7(6):1–16. http://www.health.gov.on.ca/en/pro/programs/publichealth/coronavirus/docs/operational_requirements_health_sector.pdf.

4. Rosenbaum L. The Untold Toll — The Pandemic’s Effects on Patients without Covid-19. NEJM. 2020;(382):2368–2371. doi:10.1056/NEJMms2009984

5. Markham JL, Richardson T, DePorre A, et al. Inpatient Utilization and Outcomes at Children’s Hospitals During the Early COVID-19 Pandemic. Pediatrics. 2021:e2020044735. doi:10.1542/peds.2020-044735

6. Dayal D, Gupta S, Raithatha D, Jayashree M. Missing during COVID-19 lockdown: children with new-onset type 1 diabetes (PREPRINT). Res Gate. 2020;May 13:1–7. doi:10.21203/rs.3.rs-28594/v1

7. Bryant DJ, Oo M, Damian AJ. The Rise of Adverse Childhood Experiences During the COVID-19 Pandemic. Psychol Trauma Theory, Res Pract Policy. 2020;12:193–194. doi:10.1037/tra0000711

8. Lazzerini M, Barbi E, Apicella A, Marchetti F, Cardinale F, Trobia G. Delayed Access or Provision of Care in Italy Resulting from fear of COVID-19. Lancet Child Adolesc Heal. 2020;4642(20):2019–2020. doi:10.1016/S2352-4642(20)30108-5

9. Chachlani N, Buchanan F, Gill P. Addressing the Indirect Effects of COVID-19 on the Health of Children and Young people. CMAJ. 2020:1–7. doi:10.1503/cmaj.201008

10. Wang G, Zhang Y, Zhao J, Zhang J, Jiang F. Mitigate the Effects of Home Confinement on Children during the COVID-19 Outbreak. Lancet. 2020;395:3–5. doi:10.1016/S0140-6736(20)30547-X

11. The Alliance for Child Protection in Humanitarian Action. Technical Note: Protection of Children during the Coronavirus Pandemic. 2019;(March):14–15.

12. Ontario PH. Enhanced Epidemiological Summary COVID-19 Case Fatality, Case Identification, and Attack Rates in Ontario. 2020:1–22.

13. Harris PA, Taylor R, Thielke R, Payne J, Gonzalez N, Conde JG. Research electronic data capture (REDCap)-A metadata-driven Methodology and Workflow Process for Providing Translational Research Informatics Support. J Biomed Inform. 2009;42(2):377–381. doi:10.1016/j.jbi.2008.08.010

14. Harris PA, Taylor R, Minor BL, et al. The REDCap consortium: Building an International Community of Software Platform Partners. J Biomed Inform. 2019;95(May):103208. doi:10.1016/j.jbi.2019.103208

15. Diskin C, Orkin J, Agarwal T, Parmar A, Friedman J. The Secondary Consequences of the COVID-19 Pandemic in Hospital Pediatrics. Hosp Pediatr. 2021. doi:10.1542/hpeds.2020-002477

16. Crowe S, Cresswell K, Robertson A, Huby G, Avery A, Sheikh A. The Case Study Approach. BMC Med Res Methodol. 2011;11(100). doi:10.1177/108056999305600409

17. Braun V, Clarke V. Using Thematic Analysis in Psychology. Qual Res Psychol. 2006;3(2):77–101.

18. Place R, Lee J, Howell J. Rate of Pediatric Appendiceal Perforation at a Children ‘ s Hospital During the COVID-19 Pandemic Compared With the Previous Year. JAMA Netw open. 2020;3(12):12–14. doi:10.1001/jamanetworkopen.2020.27948

19. Cherubini V, Gohil A, Addala A, et al. Unintended Consequences of Coronavirus Disease-2019: Remember General Pediatrics. J Pediatr. 2020. doi:10.1016/j.jpeds.2020.05.004

20. Offenbacher R, Knoll MA, Loeb DM. Delayed Presentations of Pediatric Solid Tumors at a Tertiary Care Hospital in the Bronx due to COVID-19. Pediatr Blood Cancer. 2020;(July):e28615. doi:10.1002/pbc.28615

21. Kuo DZ, Houtrow AJ, Arango P, Kuhlthau KA, Simmons JM, Neff JM. Family-centered care: Current applications and future directions in pediatric health care. Matern Child Health J. 2012;16(2):297–305. doi:10.1007/s10995-011-0751-7

22. American Academy of Paediatrics. Family Presence Policies for Pediatric Inpatient Settings During the COVID-19 Pandemic. Family Presence Policies for Pediatric Inpatient Settings During the COVID-19 Pandemic. Published 2020.

23. Aarthun A, Øymar KA, Akerjordet K. Parental Involvement in Decision-making about their Child’s Health care at the Hospital. Nurs Open. 2019;6(1):50–58. doi:10.1002/nop2.180

24. Boland L, Kryworuchko J, Saarimaki A, Lawson ML. Parental Decision Making Involvement and Decisional Conflict: A descriptive study. BMC Pediatr. 2017;17(1):1–8. doi:10.1186/s12887-017-0899-4

25. Kokorelias KM, Gignac MAM, Naglie G, Cameron JI. Towards a Universal Model of Family Centered Care: A Scoping Review. BMC Health Serv Res. 2019;19(1):1–11. doi:10.1186/s12913-019-4394-5

26. McGrail KM, Ahuja MA, Leaver CA. Virtual Visits and Patient-Centered Care: Results of a Patient Survey and Observational Study. J Med Internet Res. 2017;19(5):e177. doi:10.2196/jmir.7374

27. Virtual Care Playbook.; 2020. College of Family Physicians of Canada, Canadian Medical Association. Available at https://www.cma.ca/virtual-care-playbook-canadian-physicians.

28. Azzopardi-Muscat N, Sørensen K. Towards an Equitable Digital Public Health era: Promoting Equity through a Health Literacy Perspective. Eur J Public Health. 2019;29:13–17. doi:10.1093/eurpub/ckz166

29. Crawford A, Serhal E. Digital health equity and COVID-19: The Innovation Curve cannot Reinforce the Social Gradient of Health. J Med Internet Res. 2020;22(6):1–5. doi:10.2196/19361

30. Asbury K, Fox L, Deniz E, Code A, Toseeb U. How is COVID-19 Affecting the Mental Health of Children with Special Educational Needs and Disabilities and Their Families? J Autism Dev Disord. 2020;(0123456789). doi:10.1007/s10803-020-04577-2

31. Cacioppo M, Bouvier S, Bailly R, et al. Emerging Health Challenges for Children with Physical Disabilities and their Parents during the COVID-19 pandemic: The ECHO French survey. Ann Phys Rehabil Med. 2020;1428. doi:10.1016/j.rehab.2020.08.001

32. Wong CA, Ming D, Maslow G, Gifford EJ. Mitigating the Impacts of the COVID-19 Pandemic Response on At-Risk Children. Pediatrics. 2020;146(1):e20200973. doi:10.1542/peds.2020-0973

33. Baumbusch J, Lamden-Bennett SR, Lloyd JE V. The Impact of COVID-19 on British Columbia’s Children with Medical Complexity and Their Families. Vancouver : University of British Columbia Library. Available at https://open.library.ubc.ca/cIRcle/collections/facultyresearchandpublications/52383/items/1.0395118

34. Hassinger A, Lail J. Pandemic is Declared: Early Experience from Families of Children with Medical Complexity during SARS-COV-2 Lockdown: Information to Drive System Change. Complex Care J. 2021;2.

35. Shapiro J, Mc Donald TB. Supporting Clinicians during COVID-19 and Beyond -Learning from Past Failures and Envisioning New Strategies. NEJM. 2020:1489–1491. doi:10.1056/NEJMp2024834

36. Khullar D, Bond AM, Schpero WL. COVID-19 and the Financial Health of US Hospitals. JAMA - J Am Med Assoc. 2020;323(21):2127–2128. doi:10.1001/jama.2020.6269

37. Hassinger A, Lail J. PANDEMIC IS DECLARED: Early Experience from Families of Children with Medical Complexity during SARS-COV-2 Lockdown: Information to Drive System Change. Complex Care J. 2021. http://complexcarejournal.org/2021/01/05/pandemic-is-declared/.

