## Appendix for "Disruptions in Care: Consequences of the COVID-19 Pandemic in a Children’s Hospital"

**Supplementary Appendix**

**Table 1: Survey distributed to clinicians**

1. Date
2. Have you completed this survey before? If yes, proceed to question 3, If no, complete consent and demographics*.
3. Have you encountered a clinical case (non-COVID-19 positive) with a negative clinical outcome that you presume to be associated with the COVID-19 pandemic?
   This can include delayed presentation for medical attention, a consequence of health service closure or the societal changes due to pandemic mitigation (e.g. physical distancing and impact on mental health) If yes, proceed to next question. If no, survey completed
4. Please estimate the date this case presented (e.g., March 15)
5. Patient MRN
6. Provide a brief description of the pertinent details from your perspective
7. In your opinion, why do you presume that the clinical case presentation, outcome, or other component is related to the COVID-19 pandemic?
8. Do you have another case to report? If yes, repeat questions 4-7, If no, survey completed.

*(i)Position (Physician (faculty, trainee e.g. clinical fellow, subspecialty resident, core paediatric resident), dentist, advanced practice nurse), (ii) primary area of practice (division or sub-specialty), (iii)years of practice

**Table 2: Clinical Case Report form**

1. Study Id:
2. Patient age: ___years ______months
3. Gender
4. Primary Diagnosis
   a. Secondary diagnosis (if relevant)
5. Was hospital admission required: Y/ N;
   a. If no, where did the patient receive care: Emergency Department, Outpatient
   b. If yes, what service
6. Was the patient transferred from another hospital: Y/N
7. Total Length of hospital stay: ___________ days
   a. Total hospital stay at community site (if applicable): ___ days
8. Did the patient require ICU care: Y/N
   a. If yes, duration: ___days
9. Key details from admission note
   a. Duration of symptoms before presentation
   b. Contact with healthcare providers before a presentation, including:
      i. Type of contact
      ii. HCP
      iii. Frequency
   c. Clinical Management
10. Diagnostic Imaging
    a. Number of diagnostic images
    b. Type of diagnostic images
11. Key details from ICU note
    a. Time spent in ICU
    b. Invasive ventilation
    c. Inotropic support
    d. Renal replacement therapy
12. Pharmacological Management
    a. Antibiotics required
       i. Duration (days)
    b. Other
       i. Duration (days)
13. Key details from discharge note
    a. Suggested follow-up
14. Surgical intervention required: Y/N
    a. What was the surgery
    b. Was there a need for repeated interventions
15. CCRT involvement Y/N
16. Where was this case identified?
    a. Physician report
    b. ED
    c. Heads of M+M
17. Was there an identified morbidity?
18. Mortality: Y/N

| Table 3 - Clinical specialties completing survey | | |
| --- | --- | --- |
|  | N | %^‡^ |
| Department of Pediatrics | **146** | **69%** |
| Adolescent Medicine | 3 |  |
| Cardiology | 9 | 4.2 |
| Clinical Pharmacology and Toxicology | 1 |  |
| Dermatology | 1 |  |
| Developmental Medicine including Rehabilitation Medicine | 6 |  |
| Emergency Medicine | 12 | 5.6% |
| Endocrinology | 4 |  |
| Gastroenterology, Hepatology and Nutrition | 6 |  |
| Haematology and Oncology | 25 | 12% |
| Immunology | 1 |  |
| Infectious Disease | 4 |  |
| Medical Genetics | 3 |  |
| Metabolic Medicine | 2 |  |
| Neonatology | 10 | 4.7% |
| Nephrology | 7 |  |
| Neurology | 7 |  |
| Paediatric Medicine | 32 | 15% |
| Palliative Care | 2 |  |
| Respirology | 4 |  |
| Department of Perioperative Services | **51** | **24%** |
| Anesthesia and Pain Medicine | 14 | 6.6% |
| Critical Care | 7 |  |
| Dentistry | 4 |  |
| General and thoracic surgery | 2 |  |
| Neurosurgery | 4 |  |
| Ophthalmology | 8 |  |
| Orthopaedics | 5 |  |
| Otolaryngology | 3 |  |
| Plastics | 2 |  |
| Urology | 2 |  |
| Psychiatry | **4** | 2% |
| Diagnostic Imaging | **4** | 2% |
| Laboratory Medicine | **4** | **2%** |
| Clinical Medicine | 1 |  |
| Pathology | 3 |  |
| Not provided | **3** | **1%** |
